# Estimating youth diabetes risk using NHANES data and machine learning

**DOI:** 10.1101/19007872

**Authors:** Nita Vangeepuram, Bian Liu, Po-hsiang Chiu, Linhua Wang, Gaurav Pandey

**Author notes:** Current address: Baylor College of Medicine, Houston, TX, USA. Corresponding Author Information: Nita Vangeepuram, MD, MPH Address: 1 Gustave L. Levy Place Box 1077, New York, NY 10029.

## Abstract

**Background:** Prediabetes and diabetes mellitus (preDM/DM) have become alarmingly prevalent among youth in recent years. However, simple questionnaire-based screening tools to reliably assess diabetes risk are only available for adults, not youth.

**Methods:** As a first step in developing such a tool, we used a large-scale dataset from the National Health and Nutritional Examination Survey (NHANES) to examine the performance of a published pediatric clinical screening guideline in identifying youth with preDM/DM based on American Diabetes Association diagnostic biomarkers. We assessed the agreement between the clinical guideline and biomarker criteria using established evaluation measures (sensitivity, specificity, positive/negative predictive value, F-measure for the positive/negative preDM/DM classes, and Kappa). We also compared the performance of the guideline to those of machine learning (ML) based preDM/DM classifiers derived from the NHANES dataset.

**Results:** Approximately 29% of the 2858 youth in our study population had preDM/DM based on biomarker criteria. The clinical guideline had a sensitivity of 43.1% and specificity of 67.6%, positive/negative predictive values of 35.2%/74.5%, positive/negative F-measures of 38.8%/70.9%, and Kappa of 0.1 (95%CI: 0.06-0.14). The performance of the guideline varied across demographic subgroups. Some ML-based classifiers performed comparably to or better than the screening guideline, especially in identifying preDM/DM youth (p=5.23×10^−5^).

**Conclusions:** We demonstrated that a recommended pediatric clinical screening guideline did not perform well in identifying preDM/DM status among youth. Additional work is needed to develop a simple yet accurate screener for youth diabetes risk, potentially by using advanced ML methods and a wider range of clinical and behavioral health data.

**Key Messages:**

- As a first step in developing a youth diabetes risk screening tool, we used a large-scale dataset from the National Health and Nutritional Examination Survey (NHANES) to examine the performance of a published pediatric clinical screening guideline in identifying youth with prediabetes/diabetes based on American Diabetes Association diagnostic biomarkers.
- In this cross-sectional study of youth, we found that the screening guideline correctly identified 43.1% of youth with prediabetes/diabetes, the performance of the guideline varied across demographic subgroups, and machine learning based classifiers performed comparably to or better than the screening guideline in identifying youth with prediabetes/diabetes.
- Additional work is needed to develop a simple yet accurate screener for youth diabetes risk, potentially by using advanced ML methods and a wider range of clinical and behavioral health data.

## Introduction

Diabetes mellitus (DM) is a serious chronic condition associated with numerous long-term complications.(1) Prediabetes (preDM) is a precursor condition in which glucose levels are high, but not yet high enough to diagnose diabetes.(2) PreDM is reversible with lifestyle modification and weight loss, offering an avenue to avoid the adverse effects of diabetes.(2, 3) Both these conditions have become alarmingly prevalent among youth.(4, 5) According to a large prospective cohort study, an estimated 5,300 youth are diagnosed with type 2 DM annually in the US,(4) with a higher prevalence among older teens.(5) The overall prevalence of preDM among US adolescents based on nationally representative data was 17.7%, with higher rates in males (22.0%) than in females (13.2%), in non-Hispanic Blacks (21.0%) and Hispanics (22.9%) than in non-Hispanic Whites (15.1 %),(6) and in obese youth (25.7%) than in normal weight youth (16.4%).(7) Compared to adults, DM in youth is more difficult to treat (8) due to a more rapidly progressive decline in beta cell function, and an earlier onset of complications.(9, 10) The potential health and economic impact of DM is therefore even greater for youth than adults, given the greater number of years living with the disease and time to develop long-term complications.

The American Diabetes Association (ADA) has published a guideline for identifying preDM and DM among youth based on measurement of biomarkers [plasma glucose level after an overnight fast (FPG), plasma glucose level two hours after an oral glucose load (2hrPG), or hemoglobin A1c (HbA1c)].(11) In spite of this guideline, preDM is often underdiagnosed among youth.(12, 13) For example, one study found that only 1% of adolescents with prediabetes reported having been told by a physician that they had the condition.(13) In addition, despite professional consensus, many youth do not receive recommended annual checkups and preventive services.(14) Even for those in care, oral glucose tolerance testing is generally not conducted, as it requires fasting and testing over 2-3 hours, which is often challenging.(15–17) Thus, many youth with preDM/DM may be unaware of their condition, making it difficult to target the highest risk youth for prevention. A simple non-invasive, questionnaire-based screening tool is, therefore, a likely impactful first-line strategy to identify at-risk individuals before subjecting them to definitive testing and resource-intense prevention programs.(18–20)

Several such risk tools have been developed to detect the risk of prevalent (undiagnosed) and incident preDM and DM in adults.(21–24) For example, the ADA and the Centers for Disease Control and Prevention (CDC) have developed an easy-to-use patient self-assessment screener based on 7 questions to identify adults at risk for preDM and DM.(25, 26) Surprisingly, there exists no similar tool for accurately screening for preDM/DM risk among youth, despite the clinical and public health importance of these conditions. ADA published and the American Academy of Pediatrics (AAP) endorsed the only widely used clinical screening guideline for health care providers to test asymptomatic children and adolescents.(11) However, this clinical guideline has not been validated using large youth health data sets and ADA diagnostic guidelines.(11) Furthermore, such guidelines may not perform equally in different age, sex and race/ethnicity subgroups.(27)

To address these critical knowledge gaps, and as a first step in the development of a youth diabetes risk screening tool, our objective was to examine the performance of the AAP/ADA screening guideline in identifying youth with preDM/DM. Disease determination in our study was based on biomarker (FPG, 2hrPG, and HbA1c) measurements in a large-scale dataset from the National Health and Nutrition Examination Survey (NHANES).(28) We also examined how this screening guideline performed in age, sex, and racial/ethnic subgroups. Furthermore, hypothesis-free data-driven machine learning (ML) methods(29) have recently helped improve disease diagnosis, prognosis, and treatment efficacy.(30–32) Inspired by these advances, we also investigated if ML methods applied to NHANES data can help improve preDM/DM screening performance.(33)

## Methods

### Study population

We utilized publicly available data from NHANES, a large ongoing cross-sectional survey that systematically gathers data from interviews, medical examinations, and laboratory testing for studying a range of health topics.(28) NHANES oversamples certain subgroups, such as African-Americans, Hispanics, Asians, older adults, and low income populations, to obtain reliable estimates of health status indicators for these groups.

We selected 2970 youth aged 12-19 years from 2005-2016 NHANES data for which preDM/DM diagnostic biomarkers were available.(34) We excluded 112 participants that lacked information on BMI percentile, family history of diabetes, blood pressure measures or total cholesterol, making it impossible to apply the AAP/ADA screening guideline.

### PreDM/DM status

PreDM/DM status was based on current ADA biomarker criteria (elevated levels of any of the three biomarkers: FPG ≥ 100 mg/dL, 2hrPG ≥ 140 mg/dL, or HbA1C ≥ 5.7%).(11) Since few youth had DM based on biomarker diagnostic criteria (n=13), we combined youth with preDM and DM into one category. We applied the AAP/ADA screening guideline using operationally defined equivalent variables available in NHANES (**Table 1**).

**Table 1.** Pediatric clinical screening guideline used to define prediabetes/diabetes (preDM/DM) status and their corresponding operationally defined equivalent variables in the National Health and Nutrition Examination Survey (NHANES).

| **ADA/AAP preDM/DM risk for children**<br>(at-risk if overweight plus one or more additional risk factors) | **NHANES variables used**<br>(at-risk if overweight plus one or more additional risk factors) |
| --- | --- |
| Overweight (BMI >85th percentile for age and sex, weight for height >85th percentile, or weight >120% of ideal for height) **A<sup>a</sup>** | BMI ≥ 85th percentile<sup>b</sup> |
| Additional risk factors: | Additional risk factors: |
| Maternal history of gestational diabetes during the child's gestation **A<sup>a</sup>** | Not available. |
| Family history of type 2 diabetes in first- or second-degree relative **A<sup>a</sup>** | Ever been told by a doctor or other health professional that you have health conditions or a medical or family history that increases your risk for diabetes? |
| Race/Ethnicity (Native American, African American, Latino, Asian American, Pacific Islander) **A<sup>a</sup>** | Non-White race/ethnicity (non-Hispanic Black, Hispanic, other) |
| Signs of insulin resistance or conditions associated with insulin resistance (hypertension, dyslipidemia, acanthosis nigricans, polycystic ovary syndrome, or small-for-gestational-age birth weight). **B<sup>a</sup>** | Hypertension<sup>c</sup>: Blood pressure ≥90th percentile or ≥120/80 mm Hg for children ≥13 years; Dyslipidemia<sup>d</sup>: total cholesterol ≥170 mg/dL |

**Notes:** <sup>a</sup>, Evidence grades, with grade A and B representing higher and moderate quality evidence, respectively. Grades do not factor into the determination of risk in the current study.
<sup>b</sup>, We calculated BMI percentiles using the SAS program provided by the CDC for the 2000 CDC growth charts (ages 0 to <20 years) with overweight/obese defined as ≥85<sup>th</sup> percentile. Available from: https://www.cdc.gov/nccdphp/dnpao/growthcharts/resources/sas.htm.
<sup>c</sup>, We calculated blood pressure percentiles for children (<18 years) based on the 2017 Clinical Practice Guidelines from the AAP using the recommended published SAS program. Available from: https://sites.google.com/a/channing.harvard.edu/bernardrosner/pediatric-blood-press/childhood-blood-pressure.
<sup>d</sup>, We defined dyslipidemia as elevated total cholesterol level (≥170 mg/dL) according to AAP guidelines. Available from: https://www.healthychildren.org/English/healthy-living/nutrition/Pages/Cholesterol-Levels-in-Children-and-Adolescents.aspx.
<sup>e</sup>, ADA: American Diabetes Association; AAP: American Academy of Pediatrics; preDM: prediabetes; DM: diabetes; BMI: body mass index; NHANES: National Health and Nutrition Examination Survey

As a sensitivity analysis, we also used a higher threshold level in FPG and HbA1C to define preDM/DM status: FPG >110 mg/dL, 2hrPG ≥ 140 mg/dL, or HbA1C > 6.0%), as has been suggested by some organizations.(35)

### Machine learning

As alternatives to expert-defined screeners, we explored automated ML methods(29) for developing preDM/DM status (yes or no) classifiers directly from the youth NHANES data. We used the same five variables used in the AAP/ADA screening guideline, namely continuous BMI percentiles, family history of diabetes (yes/no), race ethnicity (non-Hispanic white vs otherwise), hypertension (yes/no), and continuous total cholesterol levels, as features. Ten established algorithms and a five-fold cross-validation setup were used to generate and evaluate preDM/DM classifiers from the values of these features for the youth in our dataset. Details of this classifier generation and evaluation process are provided in **Supplemental Information**.

### Evaluation of screeners

Both the AAP/ADA screening guideline, as well as the ML-based classifiers described above, produce binary classifications, specifically positive (+) and negative (-) preDM/DM determinations. Due to the inherent imbalance between these classes (**Table 3**), we used six appropriate measures(36) to evaluate these classifications: sensitivity (recall+), specificity (recall-), positive predictive value (PPV, precision+), negative predictive value (NPV, precision-), and F-measures for the two classes. **Table 3** and **Supplemental Information** provide definitions of these measures, and our detailed reasoning for focusing on them. We used the recommended Friedman and Nemenyi tests(37) to assess the statistical significance of the comparisons of the predictive performances of all the ML methods tested, as well as the screening guideline.

In the non-ML analyses, we assessed the six performance measures for the overall data and for sub-datasets stratified by sex (male, female), race/ethnicity (non-Hispanic white, non-Hispanic black, Hispanic, other), and age groups (12-14 years, 15-17 years, and 18-19 years). We examined the agreement between the AAP/ADA screener and biomarkers in defining preDM/DM using McNemar’s test and reported Kappa coefficient, which has a value ranging from 0 (no consistency) to 1 (complete consistency). We also tested equal Kappa coefficients across subgroups, and used the Breslow-Day test to examine the homogeneity of the odds ratios between preDM/DM status defined by the guideline and by biomarker measurements across subgroups. As the purpose of the current study was to evaluate the performance of the AAP/ADA screening guideline, not to make population level estimates of preDM/DM prevalence, we did not apply survey procedures to the NHANES data and reported only the unweighted results. Analyses were conducted in SAS (v9.4).

## Results

### Performance of clinical preDM/DM screening guideline

Approximately 29% of the 2858 youth in our study population were classified as having preDM/DM based on ADA/CDC biomarker criteria. The prevalence was 35.5% according to the AAP/ADA screening guideline (**Table 2**).

**Table 2.**
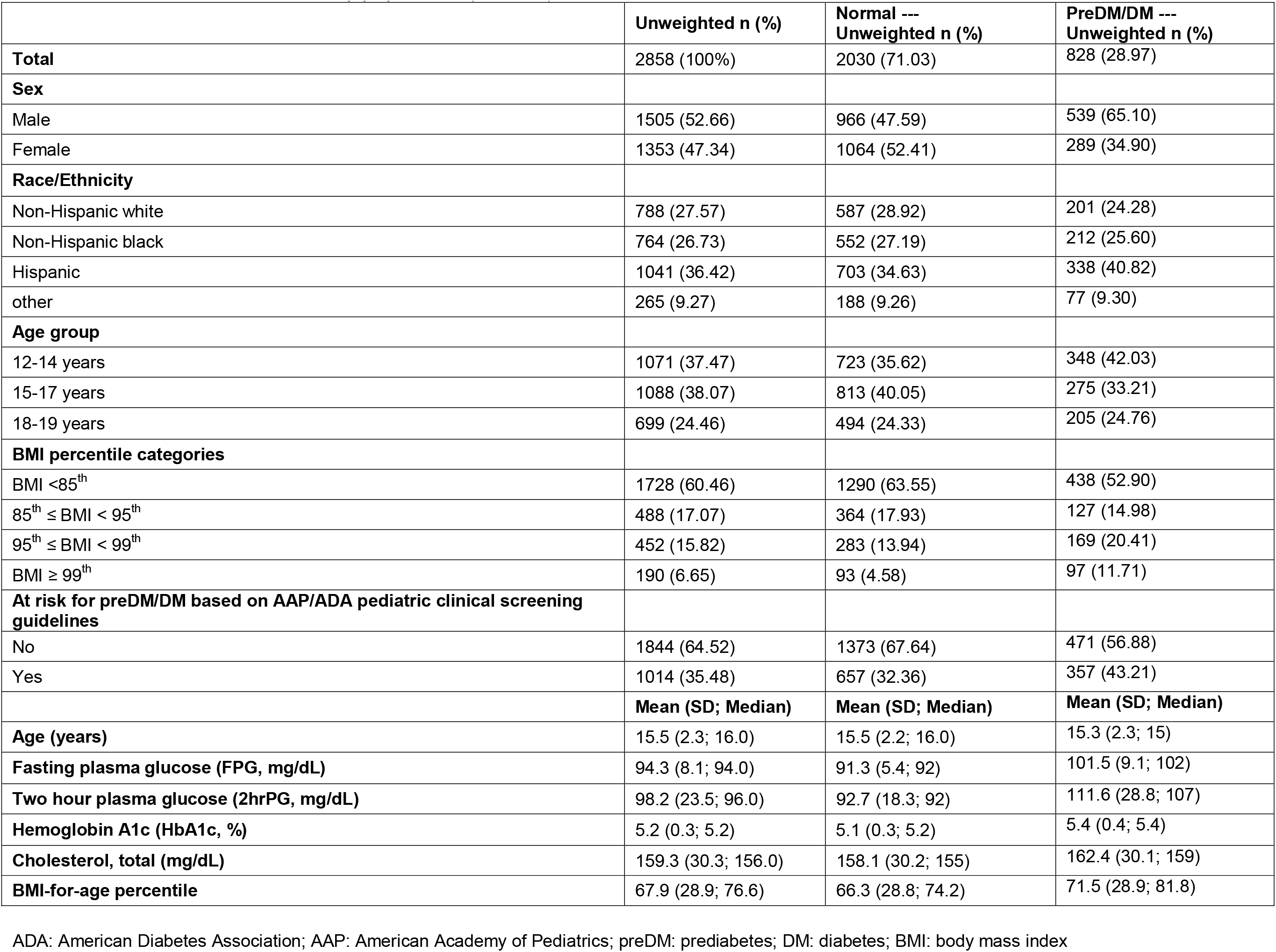
Characteristics of the study population (n=2858).

|  | Unweighted n (%) | Normal ---<br>Unweighted n (%) | PreDM/DM ---<br>Unweighted n (%) |
| --- | --- | --- | --- |
| **Total** | 2858 (100%) | 2030 (71.03) | 828 (28.97) |
| **Sex** |  |  |  |
| Male | 1505 (52.66) | 966 (47.59) | 539 (65.10) |
| Female | 1353 (47.34) | 1064 (52.41) | 289 (34.90) |
| **Race/Ethnicity** |  |  |  |
| Non-Hispanic white | 788 (27.57) | 587 (28.92) | 201 (24.28) |
| Non-Hispanic black | 764 (26.73) | 552 (27.19) | 212 (25.60) |
| Hispanic | 1041 (36.42) | 703 (34.63) | 338 (40.82) |
| other | 265 (9.27) | 188 (9.26) | 77 (9.30) |
| **Age group** |  |  |  |
| 12-14 years | 1071 (37.47) | 723 (35.62) | 348 (42.03) |
| 15-17 years | 1088 (38.07) | 813 (40.05) | 275 (33.21) |
| 18-19 years | 699 (24.46) | 494 (24.33) | 205 (24.76) |
| **BMI percentile categories** |  |  |  |
| BMI <85<sup>th</sup> | 1728 (60.46) | 1290 (63.55) | 438 (52.90) |
| 85<sup>th</sup> ≤ BMI < 95<sup>th</sup> | 488 (17.07) | 364 (17.93) | 127 (14.98) |
| 95<sup>th</sup> ≤ BMI < 99<sup>th</sup> | 452 (15.82) | 283 (13.94) | 169 (20.41) |
| BMI ≥ 99<sup>th</sup> | 190 (6.65) | 93 (4.58) | 97 (11.71) |
| **At risk for preDM/DM based on AAP/ADA pediatric clinical screening guidelines** |  |  |  |
| No | 1844 (64.52) | 1373 (67.64) | 471 (56.88) |
| Yes | 1014 (35.48) | 657 (32.36) | 357 (43.21) |
|  | **Mean (SD; Median)** | **Mean (SD; Median)** | **Mean (SD; Median)** |
| **Age (years)** | 15.5 (2.3; 16.0) | 15.5 (2.2; 16.0) | 15.3 (2.3; 15) |
| **Fasting plasma glucose (FPG, mg/dL)** | 94.3 (8.1; 94.0) | 91.3 (5.4; 92) | 101.5 (9.1; 102) |
| **Two hour plasma glucose (2hrPG, mg/dL)** | 98.2 (23.5; 96.0) | 92.7 (18.3; 92) | 111.6 (28.8; 107) |
| **Hemoglobin A1c (HbA1c, %)** | 5.2 (0.3; 5.2) | 5.1 (0.3; 5.2) | 5.4 (0.4; 5.4) |
| **Cholesterol, total (mg/dL)** | 159.3 (30.3; 156.0) | 158.1 (30.2; 155) | 162.4 (30.1; 159) |
| **BMI-for-age percentile** | 67.9 (28.9; 76.6) | 66.3 (28.8; 74.2) | 71.5 (28.9; 81.8) |

ADA: American Diabetes Association; AAP: American Academy of Pediatrics; preDM: prediabetes; DM: diabetes; BMI: body mass index

As shown in **Table 3**, the guideline correctly identified 43.1% of the youth with preDM/DM based on biomarkers (sensitivity), the PPV (precision+) was 35.2%, and the preDM/DM F-measure was 38.8%. We found poor agreement between preDM/DM determinations based on biomarkers and those based on the AAP/ADA screening guideline (Kappa coefficient 0.1 (95%CI: 0.06-0.14), p<0.0001). The Kappa coefficients did not differ by sex, age, or race/ethnicity (p>0.05), indicating that the guideline did not perform well in any of the subgroups. The agreement between preDM/DM determinations based on biomarkers and those based on the screening guideline differed between males and females (Breslow-Day test p=0.02), and across the three age groups (p=0.046). It did not differ across the four racial/ethnic groups (p=0.42).

**Table 3.** Performance measures of pediatric clinical screening guideline when compared against prediabetes/diabetes (preDM/DM) determinations based on biomarker criteria.

| preDM/DM based on elevated FPG/2hrPG/HbA1C | AAP/ADA pediatric clinical screening guidelines |  |  |  |
| --- | --- | --- | --- | --- |
|  |  | Yes | No | Row total |
|  | Yes | 357 | 471 | 828 |
|  | No | 657 | 1373 | 2030 |
|  | Column total | 1014 | 1844 | 2858 |
| **Performance measures of the adult screener/pediatric clinical screening guidelines when compared against preDM/DM based on biomarkers for the positive (+) and negative class (-)** |  |  |  |  |
| **Sensitivity (recall+)** = Proportion of at-risk based on pediatric clinical screening guidelines that have preDM/DM based on biomarkers |  | 357/828 = 43.1% |  |  |
| **Specificity (recall-)** = Proportion of not at-risk based on pediatric clinical screening guidelines that do not have preDM/DM based on biomarkers |  | 1272/2030 = 67.6% |  |  |
| **Positive Predictive Value (PPV, precision+)** = Proportion of youth identified with preDM/DM based on biomarkers among all predicted to be at-risk based on pediatric clinical screening guidelines |  | 357/1014 = 35.2% |  |  |
| **Negative Predictive Value (NPV, precision-)** = Proportion of youth not identified with preDM/DM based on biomarkers among all predicted not to be at-risk based on pediatric clinical screening guidelines |  | 1373/1844 = 74.5% |  |  |
| **F-measure+** = Harmonic (conservative) mean of Precision+ and Recall+ = $2 * (\text{Precision+} * \text{Recall+}) / (\text{Precision+} + \text{Recall+})$ | | $2 * (43.1\% * 35.2\%) / (43.1\% + 35.2\%) = 38.8\%$ | | |
| **F-measure-** = Harmonic (conservative) mean of Precision- and Recall- = $2 * (\text{Precision-} * \text{Recall-}) / (\text{Precision-} + \text{Recall-})$ | | $2 * (67.6\% * 74.5\%) / (67.6\% + 74.5\%) = 70.9\%$ | | |

ADA: American Diabetes Association; AAP: American Academy of Pediatrics; preDM: prediabetes; DM: diabetes; BMI: body mass index; FPG: fasting plasma glucose; 2hrPG: 2 hour plasma glucose; HbA1c: hemoglobin A1c

The predictive performance measures of the screening guideline also varied across the various subgroups (**Figure 1**). The sensitivity (recall+) was higher among females than males (52.2% vs 38.2%), while the PPV (precision+) was lower among females (29.4% vs 41.1%). The guideline performed better for Hispanics and non-Hispanic Blacks than for non-Hispanic Whites and other racial/ethnic groups in terms of sensitivity (51.8% and 51.9% vs 23.4% and 32.5% respectively), while the PPV was similar (28.8%-37.6%) across the four racial/ethnic groups. Finally, the guideline performed the worst for those aged 12-14 years (sensitivity=39.9%) and the best for those aged 18-19 years (sensitivity=47.8%, PPV=30.2%, and F-measure=43.7%).

**Figure 1.**
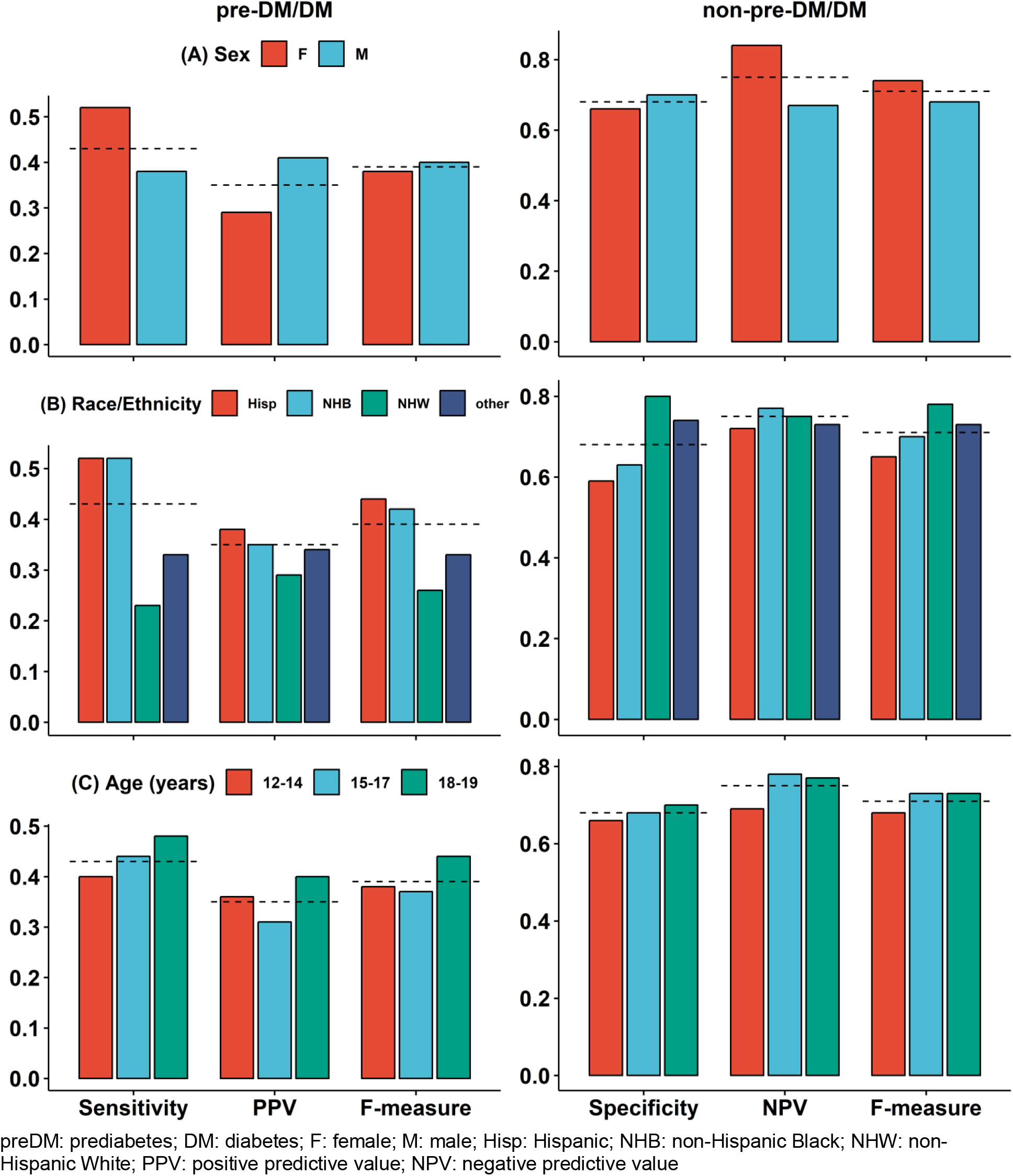
Variations in the performance of the American Diabetes Association pediatric screening guidelines in identifying youth with prediabetes/diabetes (preDM/DM) based on biomarker measurements across subgroups based on (**A**) sex (female, male), (**B**) race/ethnicity (Hispanic, non-Hispanic Black, non-Hispanic white, other) and (**C**) age. Dashed lines denote the value of the corresponding evaluation measure obtained from the full study population (youth ages 12-19, National Health and Nutrition Examination Survey data, 2005-2016).

Results from the sensitivity analysis using higher biomarker thresholds (FPG >110 mg/dL, 2hrPG ≥ 140 mg/dL, or HbA1C> 6.0%) showed similar performance measures: sensitivity=56.2%, specificity=66.0%, PPV=10.3%, NPV=95.6%, F-measure=17.3% and 78.1% for those with and without preDM/DM, respectively.

### Performance of ML-based preDM/DM classifiers

**Figure 2** shows the five-fold cross-validation(38)-derived results of classifying preDM/DM status using ML methods, variables used in the screening guideline, and class labels (preDM/DM or not) defined using biomarker criteria. Across almost all the methods and evaluation measures, it was comparatively easier to produce more accurate predictions for the bigger non-preDM/DM class than the smaller preDM/DM one. Even so, the overall performance of the ML methods varied in a manner consistent with that of the screening guideline across the evaluation measures and classes. Furthermore, in each case, at least one ML method performed better than the screening guideline, especially for the harder to predict preDM/DM class. In particular, the naïve Bayes-based classifier performed equivalently or better than the guideline in terms of all the measures for this class (Friedman-Nemenyi test p=9.216×10^-5^, 0.252 and 5.228×10^-5^ for PPV, sensitivity and F-measure respectively). This algorithm assumes conditional independence between the features, given the class labels. It then uses Bayes’ theorem to generate a simple classifier that calculates the posterior probability for a class label based on the values of the features for a given patient. The classifier based on this algorithm also performed better than or equivalently to the guideline for the non- preDM/DM class (p=8.5×10^-10^, 0.225 and 0.005 for NPV, specificity and F-measure respectively). Several other methods, such as Logistic (Regression), LogitBoost, PART and J48 (decision tree), also performed statistically equivalently or better than the screening guideline. Overall, these results show that even with very few features (only five here), data-driven ML-based methods can help improve upon the performance of the AAP/ADA preDM/DM screening guideline.

**Figure 2.**
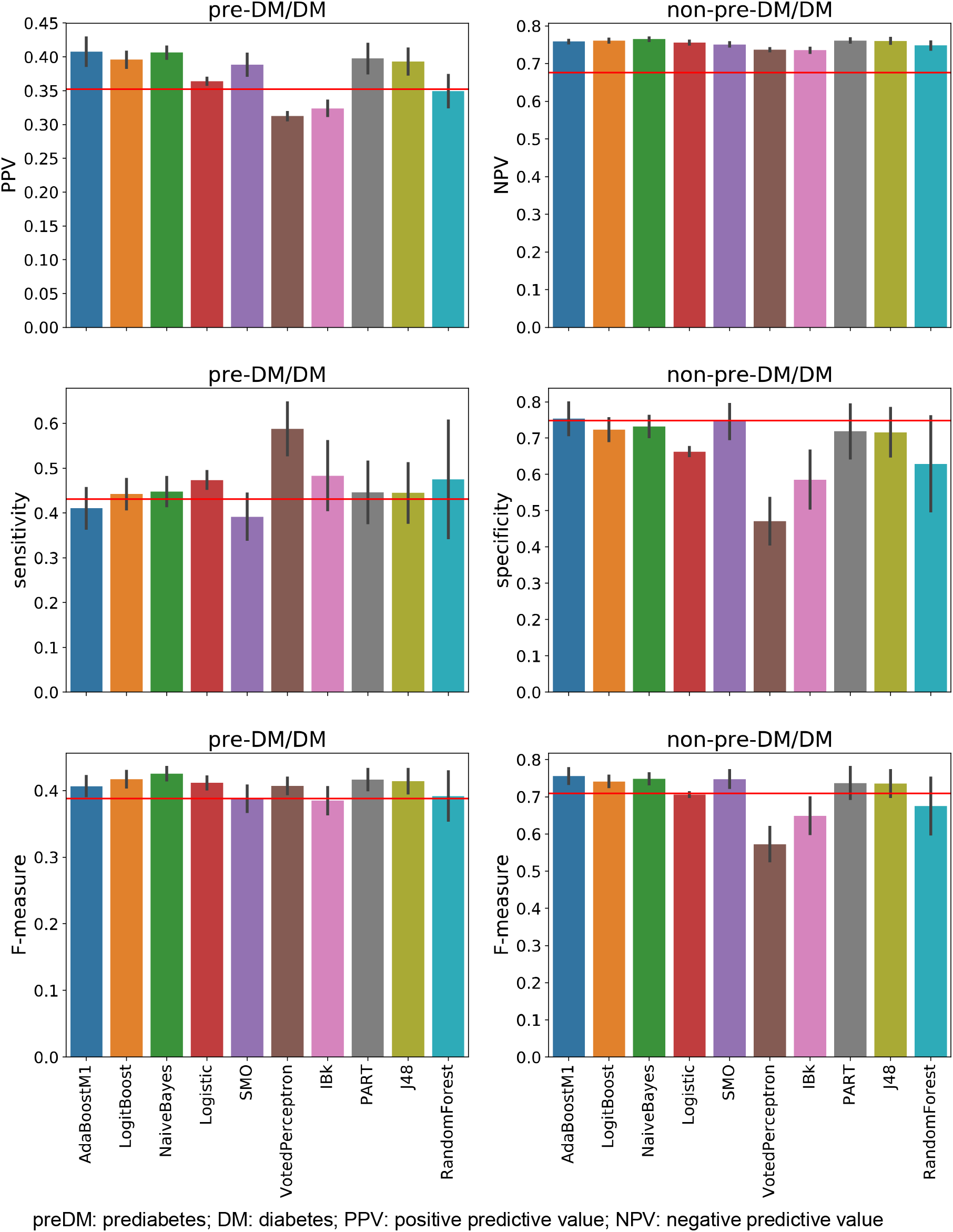
Performance of machine learning algorithms in classifying individuals into prediabetes/diabetes (preDM/DM) and non-preDM/DM classes, evaluated in terms of predictive value, sensitivity/specificity and F-measures for both classes. The variables used in this classification were the same as those used in the American Diabetes Association pediatric screening guidelines, whose performance in terms of each measure is shown by a horizontal red line in the corresponding subplot.

## Discussion

The recently increasing prevalence of preDM/DM among youth, even among those with normal weight,(7) and the underdiagnosis of these conditions despite serious long-term sequelae, point to a pressing need for the development of simple accurate screening tools for identifying at-risk youth. Towards that end, we conducted the first evaluation of a current pediatric clinical screening guideline recommended by the AAP and ADA on NHANES data, using preDM/DM status determined based on biomarker criteria (elevated FPG/2hrPG/HbA1 C) for comparison. Despite the fact that the pediatric clinical screening guideline is meant for health care providers to identify youth at risk for diabetes, the sensitivity of the guideline in identifying NHANES youth with preDM/DM based on biomarkers was below 50%. The agreement between risk based on the clinical screening guideline and presence of preDM/DM based on biomarker criteria was similarly poor across demographic subgroups based on age, sex and race/ethnicity. On the other hand, we found that the prevalence of preDM/DM varied across these subgroups, and the association between preDM/DM status defined by the guideline and based on biomarkers differed between males and females, and potentially by age groups. Another study also reported variations in the performance of diabetes risk scores by sex and race/ethnicity among adult populations in NHANES.(27) Taken together, these results suggest the need for a better screener than the current one, and a screener that can perform well for subgroup populations.

Data-driven ML-based methods(29) yielded improvements over the screening guideline in identifying youth with preDM/DM, despite using only the five variables (BMI, family history of diabetes, race/ethnicity, hypertension, and cholesterol levels) the guideline is based on. Combining many more relevant features from NHANES or other large data sets with rich clinical and behavioral health data, as well as powerful ML approaches like feature selection(39) and deep learning(40), is likely to substantially enhance our ability to develop a data-driven, relatively simple, and accurate screener for youth at risk for preDM/DM.

Of note, about half of the youth with preDM/DM in this study were of normal weight. Indeed, a recent study, also based on an examination of NHANES data, found that 16.4% of normal weight youth had preDM.(7) Another study found a relative annual increase in the incidence of type 2 diabetes, despite the fact that there was no significant increase in the prevalence of obesity among US youth in the same time period.(41) Factors other than weight status are known to increase risk of diabetes, including minority race/ethnicity and family history of diabetes.(7, 41–43) Indeed, due to their relevance, these factors are included in the pediatric screening guideline that we evaluated in our study. There are likely other factors that impact diabetes risk that are yet to be discovered. Thus, although all normal weight youth may not be at risk of developing DM, there is still value in identifying all youth with preDM, even those that aren’t obese, because they have been shown to have increased cardiovascular risk.(44) This is exactly the perspective we adopted in our study.

Despite its promising findings, our study has some limitations. PreDM/DM status was determined based on one-time measurements of biomarkers due to the data availability in NHANES, whereas the ADA recommends repeated measurements.(11) Specifically, preDM diagnosis based on a single assessment may not capture youth truly at risk for progression to DM, because preDM in adolescence is sometimes transient and related to physiologic pubertal insulin resistance.(10, 11) Furthermore, NHANES data, and thus, our evaluation, did not differentiate type 1 from type 2 diabetes. We do not expect this to substantially affect our results, since the prevalence of type 1 diabetes among youth is relatively low as compared to the combined prevalence of preDM and type 2 DM.(5, 6) Another limitation is that we were not able to exactly apply the AAP/ADA pediatric clinical screening guideline because of missing information (history of maternal gestational diabetes during the child’s gestation, presence of acanthosis nigricans, diagnosis of polycystic ovary syndrome, and history of small-for-gestational-age birthweight), or information available in a different format (family history of diabetes).

Despite these limitations, our study also has several strengths. To our knowledge, this is the first examination of the performance of a recommended pediatric clinical screening guideline for identifying preDM/DM status, determined using biomarker criteria, among youth. Our demonstration that the guideline did not perform well for this task points to the need for additional work to develop a simple yet accurate screener for youth diabetes risk. Studies focused on assessing youth preDM/DM risk to date have relied on relatively small sample sizes from localized clinical settings, and have sometimes included invasive blood tests that may not be the best initial strategy to assess risk.(45, 46) In contrast, NHANES includes a large sample of individuals from across the United States, including well-represented age, sex, and racial/ethnic subgroups, as well as detailed biomarker, clinical, and behavioral health data. While NHANES data have been used to develop diabetes risk screeners for adults,(25, 47, 48) and to examine prevalence of preDM/DM among youth,(6, 49) no studies before ours have used these data to develop and evaluate youth diabetes risk screeners. In particular, our investigation of machine learning methods applied to these data demonstrates the promise of automated data-driven methods for developing such screeners. Future work includes the use of more advanced ML methods applied to a wider range of clinical and behavioral health data available in NHANES to build better predictive tools for assessing preDM/DM risk. Such tools can be used by youth or their caretakers, as well as in clinical and community settings, to identify at-risk youth who can benefit from more intensive diabetes prevention programs.

## Data Availability

Only publicly available NHANES data were used in this study. These data are available from https://wwwn.cdc.gov/nchs/nhanes/.

## Acknowledgements

The study was enabled in part by computational resources provided by Scientific Computing at the Icahn School of Medicine at Mount Sinai.

## Author Contributions

N.V., B.L., and G.P. conceived the study and wrote the manuscript. N.V. provided clinical expertise and supervised the study. B.L. prepared the relevant NHANES data and carried out the performance analyses of the screeners. L.W. and P.C. carried out the machine learning analyses under G.P.’s supervision. All the authors reviewed and approved the manuscript.

